# Social Media Sensors to Detect Early Warnings of Influenza at Scale

**DOI:** 10.1101/2022.11.15.22282355

**Authors:** David Martín-Corral, Manuel García-Herranz, Manuel Cebrian, Esteban Moro

## Abstract

Detecting early signs of an outbreak in a viral process is challenging due to its exponential nature, yet crucial given the benefits to public health it can provide. If available, the network structure where infection happens can provide rich information about the very early stages of viral outbreaks. For example, more central nodes have been used as social network sensors in biological or informational diffusion processes to detect early contagious outbreaks. We aim to combine both approaches to detect early warnings of a biological viral process (influenza-like illness, ILI), using its informational epidemic coverage in public social media. We use a large social media dataset covering three years in a country. We demonstrate that it is possible to use highly central users on social media, more precisely high out-degree users from Twitter, as sensors to detect the early warning outbreaks of ILI in the physical world without monitoring the whole population. We also investigate other behavioral and content features that distinguish those early sensors in social media beyond centrality. While high centrality on Twitter is the most distinctive feature of sensors, they are more likely to talk about local news, language, politics, or government than the rest of the users. Our new approach could detect a better and smaller set of social sensors for epidemic outbreaks and is more operationally efficient and privacy respectful than previous ones, not requiring the collection of vast amounts of data.

## Introduction

For many viral diseases, the early detection of when and where an outbreak will appear is critical. Public administrations responsible for public health management face public health risks such as the Avian flu^1^, Zika^2^, SARS^3,4^, Ebola^5,6^ or the latest SARS-COV-2^7,8^ that can cause millions of deaths in a short period of time at global scale^9^. Traditional health surveillance systems require monitoring and detecting symptoms or case incidence in populations. However, their precision sometimes needs to be improved by the size and delayed testing methods on those populations. Combining those data sources with others about people’s mobility, the spatial spreading structure of the disease, and even other data sources seem like a promising venue to establish appropriate warning models in the early epidemic stage^10^. Novel data streams like related web search queries and web visits^11–15^, weather data^16^ or monitoring multiple digital traces at the same time^10^ have proven to be complementary and even advantageous to traditional health monitoring systems. In the same way, social media traces have been demonstrated to be a good proxy for digital epidemiological forecasting models of ILI^17–19^. Online user activity exhibits some benefits like broader spatial and demographic reach or monitoring populations that have no easy access to health services^15^.

Since some viruses are transmitted by contact on face-to-face social networks, epidemiological methods that exploit the network structure are more effective in detecting, monitoring, and forecasting contagious outbreaks^20,21^, since they allow to anticipate more accurately the transmission dynamics. Furthermore, these methods can help public health decision-makers to enhance the adoption of public health interventions^22^ like social distancing, vaccination, or behavior change campaigns, identifying those individuals most likely to get infected and spread an infectious disease or behavior (e.g., super-spreaders), or which places are more likely to be visited by those individuals^23^. This allows more efficient vaccination campaigns^24^ when the vaccination of an entire population is not possible or recommended.

The key idea behind using high-connected individuals to monitor epidemic spreading is that they are more likely to be reached by the infection. In general, human social sensing, when carefully selected, can help predict and explain social dynamics better^25–27^. In the absence of complete detailed data about contact networks, simple approaches like the friendship paradox^28^ can be used to identify more connected and central individuals (sensors) in the network that can give early signals and anticipate the spreading of information, behavior or disease before it reaches a significant fraction of the population. In particular, the friendship paradox has already been found advantageous to identify sensors for detecting influenza^29–31^ or COVID-19^32^. In social media, a previous study demonstrated the detection of global-scale viral outbreaks of information diffusion^33^ by monitoring high-degree users on Twitter.

In this work, we address the question of how we can use sensors for information propagation in online social media to get better early warning signals of a biological epidemic. We hypothesize that social media connectivity and activity are a proxy of social interactions in the real world. Thus, highly-connected users in social media (online sensors) also mirror highly-connected individuals (offline sensors) in the physical contact network. This hypothesis is based on the wealth of literature showing that online networks mimic offline contacts’ connections, similarity, and spatial organization^25,34,35^. Furthermore, we study if it is possible to identify better social media sensors automatically based on their centrality (degree) and mobility, and content behavior. We found that social media sensors can serve as early warning predictors of the exponential growth of an epidemic several weeks before the peak. The current global pandemic threads make it vital to improve the efficiency of Early Warning Epidemiological Systems (EWES) by using operationally efficient methods to anticipate the exponential growth of a virus in a community, region, or country without compromising the citizens’ privacy. Our method provides such a system in a fully privacy-preserving framework.

## Results

We used social media traces obtained from the micro-blogging site Twitter, where we collected more than 250 million tweets from December 2012 to April 2015 on Spain’s mainland. Using Natural Language processing techniques, we only included first-person ILI-related posts, summing up a population of 19696 users with at least one first-person ILI-related mention, which comprised a total of 23975 tweets (*Methods & SM Appendix* section 1 discusses our methodology). We also made use of official ILI cases from the surveillance system for influenza in Spain (ScVGE)^36^ managed by the Instituto Carlos III de Salud^36^. This system reported weekly ILI cases in Spain for each province with two weeks of delay in the state of the seasonal flu epidemic based on the current European Union proposal that regulates ILI surveillance^37^. Our dataset of official ILI cases ranges from December 2012 to April 2015 and includes three different seasons of influenza outbreaks in Spain.

Figure 1 shows a generalized ILI season from the average of ILI cases and ILI-related mentions for the three seasons. ILI cases and ILI-related mentions time series have a Pearson correlation of 0.87 (CI [0.79, 0.93] and *p*_*value*_ *<* 0.001). Since different outbreaks happen at different times of the year, we have shifted each influenza outbreak to the time of its peak. We can see that ILI-related mentions precede the official ILI cases at the beginning of the growth stages before the peak. Previous studies have proved this^17–19^. Mentions of the outbreak in social media seem to precede the exponential growth in the total population. ILI-related posts peak at -15 weeks could be related to the start of the cold season and users mixing ILI symptoms with cold symptoms, stating that they are suffering from ILI. We found a similar pattern in Google trends data.

**Figure 1.**
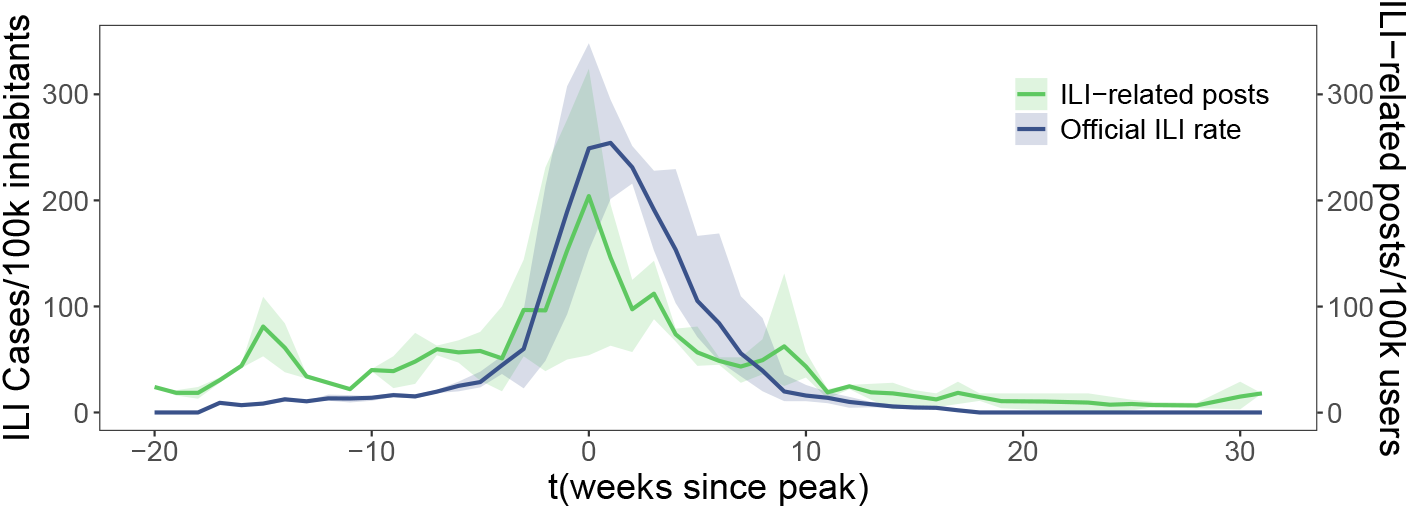
Dynamics of an average ILI season. Average ILI prevalence and ILI-related posts on Twitter across three seasons from December 2012 to April 2015 in Spain. Time (in weeks) is centered to the peak for each season. Lines are average weekly incidences for Official ILI rates (Blue) from Instituto Carlos III de Salud and first-person ILI-related mentions rate (Green) from Twitter. Bands are their confidence intervals.

### Validating high-degree individuals as sensors

However, here, we want to go a step further. Can we subset the users posting ILI-related posts to get better earlier warnings about the outbreak than monitoring the whole social network platform? Similarly to^29^, and^33^, high-degree users could be better than the average individual on the platform. To test whether high centrality or degree correlates with early signals, we measure the total weekly out-degree, *D*_*t*_, of users having social ILI-related mentions before and after the peak. Figure 2 shows distributions for *D*_*t*_ before the peak, after the peak and for the whole season. There is a statistically significant difference in the mean (*p*_*value*_ *<* 0.01). The average total weekly out-degree is 31108 (Confidence Interval, CI [21539.03, 40677.32]) before the peak, while it is only 14373 (CI [11202.94, 18455.78]) after the peak. The difference is also present in extreme values. We modelled large values of *D*_*t*_ as power laws with an exponent of 2.56 (CI [2.51, 2.62]) for the whole period. For the weeks before the peak, it follows an exponent of 2.10 (CI [1.91, 2.29]). Finally, for the weeks after the peak, it follows an exponent of 2.86 (CI [2.48, 3.25]). Thus, on the aggregated level, we indeed see that the users in social media that have ILI mentions before the peak have more social connections than after the peak. This result signals the possibility of using high-connected users as potential early sensors. This result is robust against other aggregated degree centrality variables (see *SM Appendix*, section 2). For selecting sensors, we selected each individual with an out-degree greater that 1000 (see *SM Appendix*, section 3).

**Figure 2.**
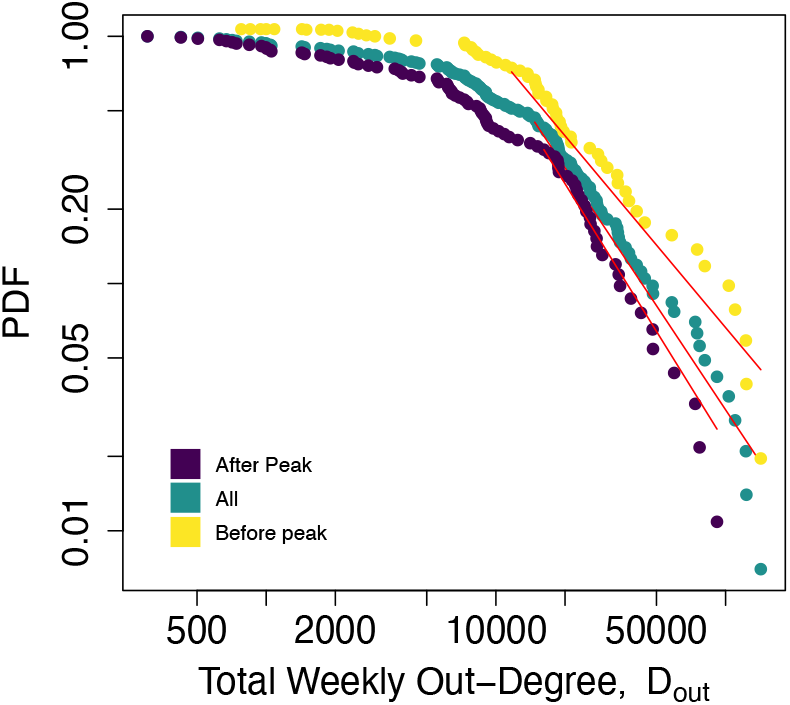
Total weekly out-degree before and after the peak. Total weekly out-degree, *D*_*t*_, power law distributions for the whole season (Green), weeks before (Yellow) peak and weeks after peak groups (Purple). Horizontal axis, total weekly out-degree, *D*_*t*_. Vertical axis probability distribution functions.

Figure 3.A compares Twitter’s cumulative ILI-related mentions of our control and sensor groups against the official ILI-related cases. As we said before, the activity in social media for both the control and sensor groups anticipates the cumulative incidence of ILI cases by one or two weeks. For each user *i* we define 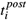 as the time in which she has an ILI-related post on social media. Figure 3.B shows confidence intervals for ILI-related posting times for each group and ILI season, relative to the peak 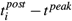. For all ILI seasons, the control group has an average ILI-related posting time of 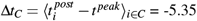 (CI [−5.54,−5.17]) weeks before the peak. The sensor group has an average ILI-related posting time of 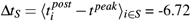 (CI [−7.42,−6.02]) weeks before the peak. This yields that sensors are posting on average Δ*t*_*S*_ Δ*t*_*C*_ = 1.37 (CI [−2.08,−0.64] and *p*_*value*_ *<* 0.01) weeks before the control group, during the exponential growth phase, between 8 to 4 weeks for all seasons. In more detail, the 2012-2013 season has a Δ*t*_*S*_ Δ*t*_*C*_ = 0.62 (CI [−1.58,−0.84] and *p*_*value*_ *>* 0.1), the 2013-2014 season has a Δ*t*_*S*_ Δ*t*_*C*_ = 2.46 (CI [−3.45,−0.36] and *p*_*value*_ *<* 0.01) and the 2014-2015 season has a Δ*t*_*S*_ Δ*t*_*C*_ = 1.54 (CI [−2.45,−0.63] and *p*_*value*_ *<* 0.01). As we can see, the ILI-related mentions of sensors could anticipate the epidemic’s growth by 1 or 2 weeks with respect to other users in the platform.

**Figure 3.**
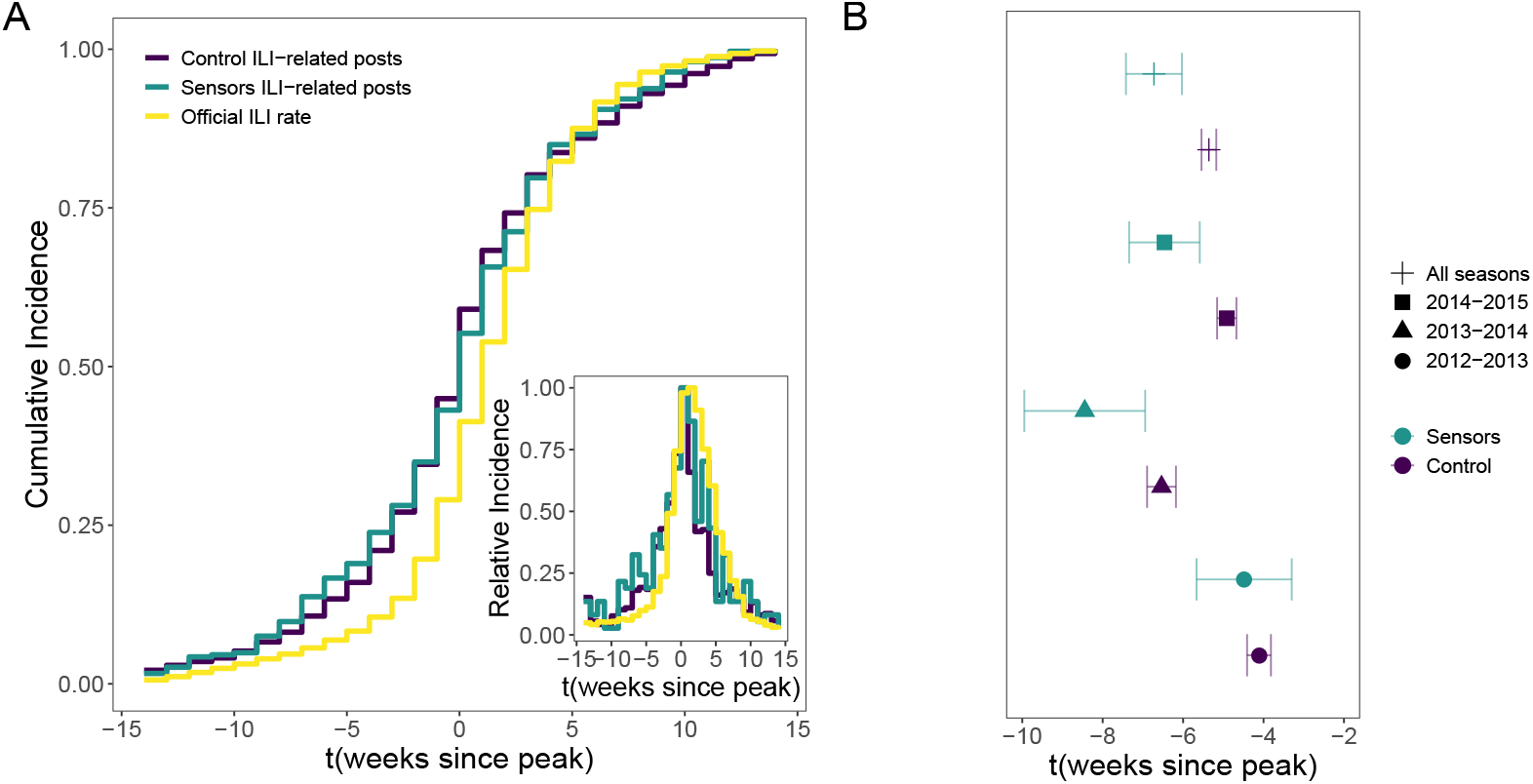
Cumulative incidence between real ILI, all Twitter, and only Twitter sensors. (A) Empirical cumulative distribution differences in official ILI cases (Yellow), control ILI-related mentions on Twitter (Purple), and sensor ILI-related mentions on Twitter (Green). Horizontal axis measures weeks since the peak on ILI cases. Inset. weekly incidence for ILI cases, control ILI-related mentions and sensor ILI-related mentions on Twitter. (B) Confidence intervals for ILI-related posting times relative to the peak 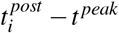 for control (Purple) and sensors (Green) groups for each ILI season. 2012-2013 (Circle), 2013-2014 (Triangle), 2014-2015 (Square) and all seasons (Cross).

### Autoregressive models with sensors and its theoretical validation

To quantify statistically how valid our sensors in social media could be in a potential EWES model, we built an autoregressive model that considered different epidemiological and social media features (see *Methods* section). The models considered different combinations of the total number of weekly ILI cases at time *t, I*_*t*_, the total weekly out-degree of all users from the social media platform (*D*_*T,t*_) that posted ILI-related mentions, and the total weekly out-degree of the subset of those users in the sensor group (*D*_*S,t*_). We have also considered different temporal week lags, *t*−*δ*, for each variable to test their potential role as early warning signals. As a baseline, we have considered a model that only incorporates the ILI cases and their autoregressive power at *t* − 1. As we see in Table 1, that simple model is already quite accurate in explaining the evolution of the weekly ILI rate. On top of that baseline model, we built four others, including the degree centrality of all users and the sensor group at different lags. For each model, we predict the *I*_*t*_ number of ILI-related cases using the information of the *I*_*t*−1_ cases and the total out-degree of all users and sensors with ILI-related mentions at time *t* and *t*−*δ*. We ran all models using a step-wise approach to keep only statistically significant regressors for *δ* = 1, 2, 3, 4. Due to multicollinearity problems between variables, we also monitor the variance inflation factor (VIF) for each to choose the best *δ*. Results in Table 1 and Figure 4A quantitatively show the importance of social media ILI-related mentions, especially those from the sensor group. As we can see, the predicting power (adjusted *R*^2^) on next week’s official ILI rate after incorporating social media mentions increases significantly (and we also reduced collinearity), especially at three-or four-week lags. In all those cases, the total degree of sensors at time *T* and time *t*−*δ* has a significant regression coefficient and role (in *R*^2^) in the prediction. That is, social sensors can help anticipate official ILI cases three to four weeks before, a result consistent with previous similar analyses of ILI contagious outbreaks in small settings^29^ or of information spreading in social media^33^. We also note that the signs of the variables of all users and sensors have different effects. For example, a higher total degree of sensors at times *t*−*δ* predicts more ILI-related cases (positive coefficient) at time *t* for *δ >* 0, but a smaller number of cases (negative coefficient) for *δ* = 0. As we will see below, this apparent contradiction comes from the high auto-correlation of the time series of ILI-related cases and the total degree of users.

**Table 1.** Empirical ILI regression models. Regression table with normalized beta coefficients for each group of variables, Official (I)LI, (T)witter and (S)ensors, where *X*_*t*_ are weekly ILI related variables for each group. *D*_*T,t*_ and *D*_*S,t*_ are weekly total out-degree variables from Twitter (T) and Sensors (S).

| | Official weekly ILI rate, $I_t$ | | | | |
| --- | --- | --- | --- | --- | --- |
| | I | I+T+S<br>$\delta = t - 1$ | I+T+S<br>$\delta = t - 2$ | I+T+S<br>$\delta = t - 3$ | I+T+ S<br>$\delta = t - 4$ |
|  | (1) | (2) | (3) | (4) | (5) |
| $I_{t-1}$ | 0.924***<br>(0.041) | 0.789***<br>(0.047) | 0.856***<br>(0.049) | 0.849***<br>(0.045) | 0.800***<br>(0.044) |
| $D_{T,t}$ | | 0.717***<br>(0.0003) | 0.634***<br>(0.0002) | 0.580***<br>(0.0002) | 0.561***<br>(0.0002) |
| $D_{T,t-1}$ | | -0.281***<br>(0.0003) | | | |
| $D_{T,t-2}$ | | | -0.344***<br>(0.0003) | | |
| $D_{T,t-3}$ | | | | -0.313***<br>(0.0002) | |
| $D_{T,t-4}$ | | | | | -0.217***<br>(0.0002) |
| $D_{S,t}$ | | -0.443***<br>(0.0004) | -0.393***<br>(0.0003) | -0.348***<br>(0.0003) | -0.339***<br>(0.0003) |
| $D_{S,t-1}$ | | 0.211***<br>(0.0005) | | | |
| $D_{S,t-2}$ | | | 0.227***<br>(0.0004) | | |
| $D_{S,t-3}$ | | | | 0.186***<br>(0.0003) | |
| $D_{S,t-4}$ | | | | | 0.132***<br>(0.0003) |
| Constant | 0.000<br>(4.627) | 0.000<br>(4.093) | 0.000<br>(4.071) | 0.000<br>(4.123) | 0.000<br>(4.516) |
| Observations | 87 | 87 | 86 | 85 | 84 |
| $R^2$ | 0.854 | 0.925 | 0.932 | 0.935 | 0.929 |
| Adjusted $R^2$ | 0.852 | 0.920 | 0.928 | 0.931 | 0.924 |
| Maximum VIF | NA | 15.16 | 9.36 | 6.77 | **5.28** |
| Residual Std. Error | 34.092 | 25.042 | 23.951 | 23.529 | 24.741 |
| F Statistic | 497.040*** | 199.556*** | 218.885*** | 226.755*** | 203.163*** |
| *Note:* *p<0.1; **p<0.05; ***p<0.01 |  |  |  |  |  |

**Figure 4.**
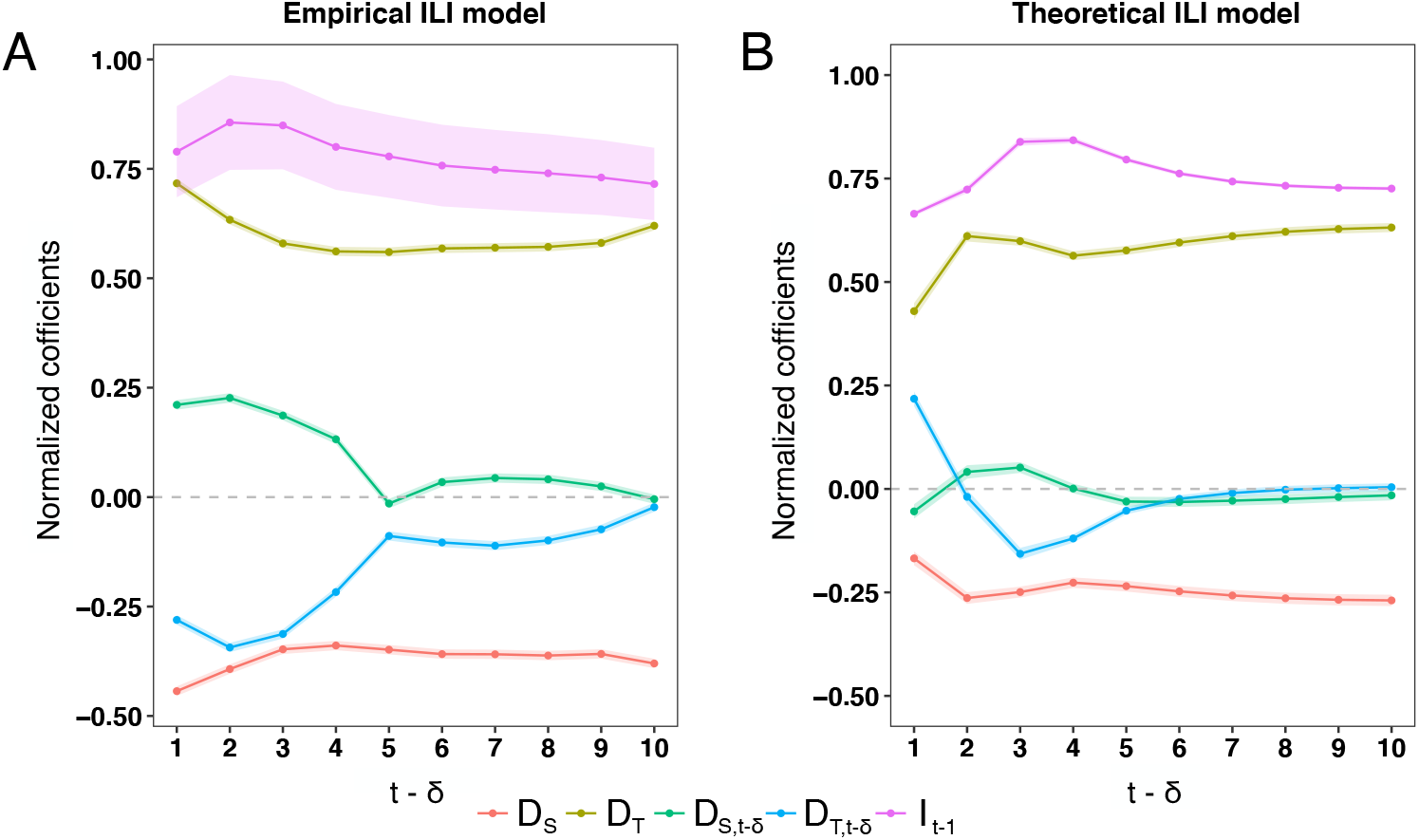
Results for the Empirical and Theoretical ILI auto-regression models. (A) Normalized coefficients for the different autoregressive models for *I*_*t*_, see Eq. (4) for different time lags *δ*. Model regressors for each *δ* are the number of cases one week in the past *I*_*t*−1_, the total out-degree at time t, *D*_*T,t*_, the total out-degree at time *t*−*δ, D*_*T,t*−*δ*_, and the total out degree of sensors at time *t, D*_*S,t*_ and at time *t*−*δ, D*_*S,t*−*δ*_. We show the normalized coefficient and their confidence intervals (shaded area). (B) Same as in (A) but for the agent-based model of ILI disease and information diffusion.

We investigated the predicting power of high-degree sensors in a synthetic model to validate that sensors anticipate ILI cases because social media connectivity mirrors social connections in the real world. Specifically, we built a base agent-based susceptible-infected-recovery (SIR) epidemic spreading on a random network mimicking real (face-to-face) social contacts between people (see *Methods* for details about the network and simulations details). Apart from their physical contacts, we also assumed that each person has acted on a social media platform and that the degree in both the real and online networks are correlated moderately. Assuming that agents post on social media when they are infected, we also constructed the time series 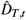 and 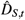 for the model and their autoregressive fits as in Table 1. Our results once again show that high-degree agents (sensors) carry some predicting power on the epidemic spreading.

Furthermore, the coefficients for the different models show the same regression structure as the empirical models in 1, see Figure 4A. We can see that both coefficient structures are nearly the same, including their magnitude and signs. Although this is not direct proof of our hypothesis that the online and offline centrality of real users is similar, it shows that under that assumption, we not only get that the effect of sensors is the same as we found in our empirical analysis, but even the structure of coefficients (magnitude and sign) is similar. These results support the idea that sensors in an informational epidemic that mirrors a biological epidemic are also sensors of a biological epidemic, like ILI, that we can trace on Twitter.

### Identification of sensors beyond out-degree

So far, we have seen that high out-degree users in social media can be early sensors of ILI cases. However, can we identify a better group of sensors beyond high degrees by looking at other traits? Are individuals that signal the epidemic’s early stages defined just by their centrality degree, or do they have other behavioral or content traits? To do that, we define a sensor functionally as every user who posts an ILI-related tweet from fifteen weeks to two weeks before the epidemic’s peak (−15 ≤ *t* ≤ −2). On the other hand, a control user was a random user who did not talk about ILI during the same period. (see *Methods* contextual features for more details).

To characterize users’ content, behavior, and network traits in both groups, we analyzed every tweet they posted 30 days before their first ILI-related tweet (sensors) or a randomly chosen tweet (control). Specifically, we identify three groups of traits for each user. Firstly, we extract the content of each user’s tweets and classify them into topics like sports, politics, entertainment and many other categories using the TextRazor classifier (see *Methods*). Secondly, since our tweets are geolocalized, we extract the mobility features of each user, in particular, the radius of gyration, which measures the size of the area covered while moving around^38^. The radius of gyration could proxy the number of different and diverse people the user is in daily contact. Thus it might serve to estimate potential exposure to infected people^39^. Lastly, we also use their activity (number of posts) and, as before, their out-degree in the social network.

To test how relevant those groups of traits are to define a sensor, we developed a straightforward logistic regression model (see *Methods*) to classify users into the sensor or control groups using different variables. As we can see in Figure 5, the accuracy of our models is above the primary level (0.5). While Network and Content groups independently achieve similar accuracies (∼0.61) than the Mobility group (∼0.62), we get better accuracy, including all types of traits (∼0.64). This result signals that even different traits carry complementary information about who could be sensors in the social media platform. To understand this further, we looked into each trait’s (normalized) coefficients in our model. As shown in Figure 4A, the most crucial variable to predict a user in the sensor group is still the out-degree in the social network, even after controlling for the number of posts. This is important because it shows that our simple method of using high-connected Twitter users as sensors works much better than other traits. We also see a small but significant effect on the radius of gyration, meaning, all things equal, users that move further are more likely to be sensors. Regarding the content, we see a structure of topics that users in the sensor group are more likely to discuss, like National, Language, Politics, and Government. On the contrary, users that talk about Sports, Popular topics, or Entertainment are less likely to be in the sensor group. This finding could signal and be related to other unobserved user traits like income or educational attainment level, which also are known to be related to the activity in social media^40^ and amount of real offline contacts^41^.

**Figure 5.**
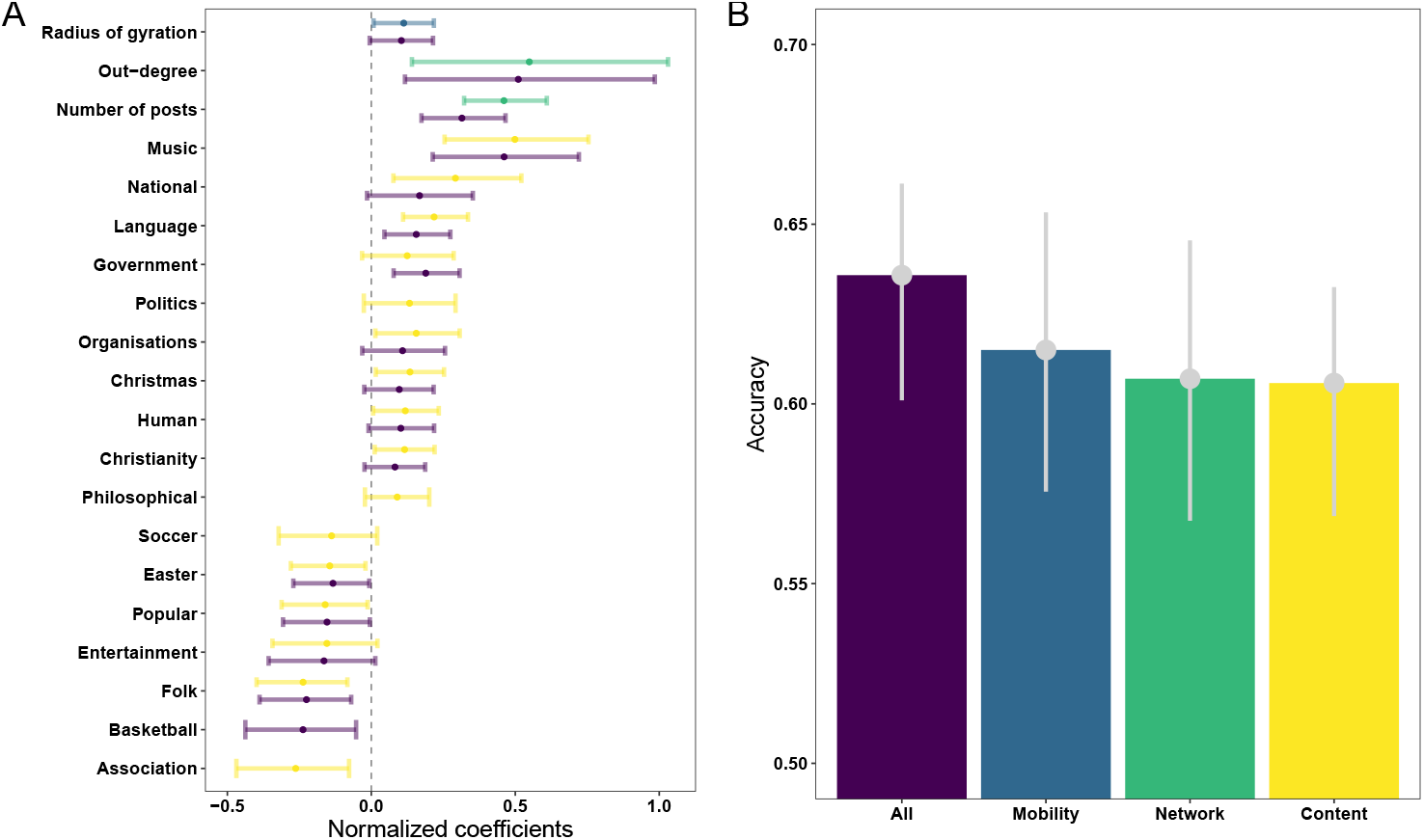
Detecting better sensors. (A) Normalized *β* coefficients from logistic models [see Eq. (7)] for each group of features for explaining sensors: content topics they posted (yellow), their network features (green), their mobility by the radius of gyration (blue) and all group variables together (purple). (B) Accuracy metrics for each group of variables: topics (yellow), network (green), mobility (blue), and all variables (purple).

## Discussion

Early warning epidemiological systems (EWES) detect outbreaks weeks in advance to help public health decision-makers make more efficient allocations of public resources to avoid or minimize an overflood of contagious in the healthcare system. EWES are undergoing significant investments and changes due to the COVID-19 disruption. However, most of them harvest vast amounts of data and do not exploit the explanatory and predictive power of the network heterogeneity where a disease-informational epidemic is spreading.

In this study, we demonstrated that social media traces, like Twitter, could be used as a source of social-behavioral data to monitor disease-informational epidemics that mirror offline biological contagious disease epidemics, like ILI, by exploiting the network heterogeneity whenever social centrality measures of the network are available. By having a simple centrality metric, such as the out-degree, we can define suitable sensors for the disease-informational epidemic in the network. When aggregated correctly, we can use sensors to feed autoregressive models that could yield signals of an outbreak up to four weeks in advance. Although previous studies showed the advantage of using social network metrics to detect, monitor, and forecast contagious outbreaks^20,21^. The usage of sensors in a network to detect early warnings of an outbreak in a biological disease contagious epidemic^29,30^, or informational epidemics^33^, our study is the first to combine the use of sensors in social media to anticipate epidemics in real life. Our results are based on the hypothesis that social media networks are related to offline contact networks, which has been validated directly in other works^25,34,35^. Our empirical and theoretical results show that instead of harvesting large amounts of data and metrics from social networks^19^, we can track and anticipate early outbreaks of a disease-informational epidemic by inexpensively looking at a small set of specific users (sensors).

We also demonstrated that sensors could be profiled and detected automatically from social media raw data by using their topological network properties and based on the content posted by individuals and their mobility patterns. Explicitly, we found that sensors talk more about some topics like National, Politics, and Government and less about Sports and Entertainment. The fact that those topics could also be related to their income, educational attainment^40^, but also to other traits like more extroversion personality traits^42^ opens the possibility to investigate the potential overlapping reasons why sensors not only are more prone to get infected earlier but also that they would like to post about it on social media. For instance, the Music topic requires further investigation; previous literature suggests individual differences in personality in the way we use and experience music^43^, possibly having a social component.

Finally, our method uses the out-degree in the social media platform as a proxy for centrality. Better knowledge of the network structure could yield more optimized methods to detect highly-central users. Our approach also has other limitations. For example, our data corresponded only to a given epidemic in a given country and were not tested against more global epidemics like the COVID-19 pandemic. However, given that our findings rely on the collective behavior of people in social media and the observed relationship between offline and online networks^44,45^, we think that our findings could be extrapolated to other epidemiological situations. We hope our research can help study the role of sensors in other pandemics, specially COVID-19, where more information about real-world offline contact networks exists due to better mobility data^46^ or contact tracing applications.

In summary, this study proposes a feasible approach to exploit the network heterogeneity underneath social media sites, like Twitter, to detect more efficiently and earlier outbreaks from a disease-informational epidemic that mirror a biological disease contagious epidemic, like ILI. Furthermore, the sensors approach we used to detect early outbreaks within informational epidemics and biological contagious disease epidemics, but this is the first time in a disease-informational epidemic as we have done in this study. Finally, novel epidemiological systems have been developed for other pathogens such as Zika, SAR, or COVID-19, among others, in addition to influenza, using conventional and non-conventional data sources such as the official public cases, online searches, or health forums. For instance, for the COVID-19 pandemic, some studies used social media traces to try to predict the dynamics of the pandemic^47,48^. Such approaches, along with our findings about the power of the network structure, could improve the results of their predictions.

Also, health systems and health organizations initiatives, like the Global Outbreak Alert and Response Network (GOARN)^49^ from WHO that is composed of 250 technical institutions and networks globally and projects like the Integrated Outbreak Analytics (IOA)^50^, Epidemic Intelligence from Open Sources (EIOS)^51^, and Epi-Brain^52^ that respond to acute public health events. This network is already moving in a double direction of incorporating early warnings from Big Data, social sciences techniques and behavioral data into epidemic response systems^53^ to control outbreaks and public health emergencies across the globe. Also, syndromic surveillance platforms like InfluenzaNet could ask for Twitter profiles or the number of people an individual interacted with in the last week to reweight the impact of different users in the prediction. Our innovative approach might help detect early outbreaks without having to monitor and harvest data from a whole population, making EWES more accurate in time prediction of an outbreak, more efficient in resources, and more respectful regarding citizens’ data privacy.

## Methods

### Data collection

We extracted Twitter data through their streaming API^54^ that allowed us to collect data programmatically on the Spanish mainland. The official ILI rate data was extracted through a web crawler built ad-hoc for the web of the Institute Carlos III of Health since there was no access to the raw data from an open data portal or a programmatic interface.

### ILI-related keywords based search and tweets classification

To get ILI-related mentions from users in the social media platform, we first filtered tweets by keeping those that mentioned simple terms like “flu“ or other ILI-related words (see SM Appendix). After that, we only kept first-person ILI-related mentions to exclude general or not directly-related posts like ‘The Spanish flu was an unusually deadly influenza pandemic’. This was done using Natural Language Processing methods. We applied a text classifier using a `scikit-learn` implementation^55^. We handpicked and labeled a set of 7836 tweets to train our classifier, containing 3918 true positive (first-person) tweets and 3918 true negative tweets. Using that labeled data, our classifier achieved an accuracy of (∼0.94). We then applied our classifier to identify first-person mentions in the remaining tweets (see SM Appendix for more details about our pipeline). After this process, we ended up with *N* = 19696 users and 23975 tweets classified as first-person ILI-related posts.

### ILI-related post time series

We added up and normalized the number of weekly users mentioning the flu by the total number of users in the system. We followed equation

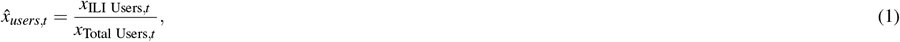

where *t* is the week. This time series is shown in Figure 1, together with the prevalence of ILI cases.

### Centrality features

Each tweet at time *t* has information about the out-degree (followees), *d*_*out,i,t*_, and in-degree (followers), *d*_*in,i,t*_, for each Twitter user *i* posting it. We used them as proxies of the network centrality for each user. Only 5% of used have more than one ILI-related mention and their in and out degrees do not change dramatically so we take *d*_*out,i,t*_ ≃ *d*_*out,i*_ (similarly for *d*_*in,i,t*_) with *t* being their first (or most of the times only) ILI-related mention. We tested out several aggregated centrality features for the selection of sensors. We calculated the weekly total, mean, median, maximum and minimum out-degree of individuals before and after the peak making first-person ILI-related mentions to test which centrality metric had more explanatory power. We found that the weekly total out-degree was the best centrality metric to apply. See *SM Appendix*, section 2 for further details.

The weekly total out-degree is defined by

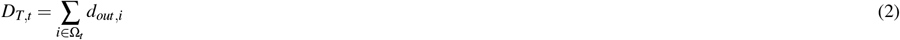

where Ω_*t*_ is the set of users that made an ILI-related mention at week *t*.

Sensors are selected as the group of users with *d*_*out,i*_ *>* 1000. For that group, we also define the time series of their centrality as

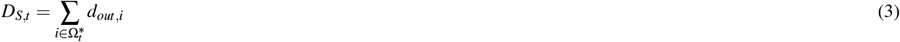

where 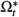 is the set of users in the sensor group that made an ILI-related mention at week *t*.

### Linear autoregressive model

The following equation represents a linear autoregressive model for explaining and nowcasting the dependent variable, *I*_*t*_, being the Official ILI rate for each week. *D*_*T,t*_ are total weekly out-degree for the whole twitter population, and *D*_*S,t*_, are total weekly out-degree for the whole sensor population. We followed

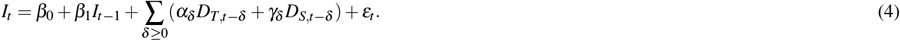

### Agent-based model of ILI disease and information diffusion

To understand our empirical findings, we compare them with the simulations of epidemic spreading on a physical and online network through an agent-based model (ABM). We model the offline (physical) contacts using a random heavy-tailed network. Specifically, we created a synthetic population of *N* = 150*k* agents with are connected through a scale-free network with degree distribution *P*(*k*) ∼ *k*^−3^ obtained through the Barabasi-Albert model.^56^. The network was built using the R package `igraph`^57^.

At the same time, we supposed that each agent participates in a social media platform. We hypothesize that the online degree of the agents is related to the offline degree in the complex network. To account for some variability, we assumed that the degree in the social media platform was modified by a random uniform distributed number (See *SM Appendix* section 4 for more details). Thus, the degree in the social media platform is given by 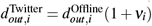, where *ν*_*i*_ is a random number uniformly distributed between 0 and 1. This way we account for potential variability between offline and online degrees.

We simulate the ILI spreading using a simple Susceptible-Infected-Recovered (SIR) epidemic model. In particular, at each time-step *t*, the infectious (I) agents can transmit the disease to their susceptible (S) neighbors in the contact network with probability *β*. If the transmission is successful, the susceptible node will move to the (I) state. An individual will move independently to the recovery (R) state with a probability *α*. We initialized the model with two initial infected seeds. After getting infected, we assumed that the agent immediately posted an ILI-related tweet on the social media platform. In our model, we considered a user to be sensors if she has an out-degree in the platform higher than four times the average degree in the Barabasi-Albert model. We also calibrated the time unit in this model so that the epidemic curves have a similar time scale as the real ILI rate (See *SM Appendix* section 4 for further details on the simulation’s parameters).

### User traits

To characterize the different traits of Twitter users, we analyzed each user’s tweets during a time window of 30 days before the initial event. For the sensor group, we selected individuals with an out-degree *d*_*out,i*_ ≥ 1000 and that made at least an ILI-related mention during the weeks −15 ≤ *t* ≤ −2 before the peak of the epidemic. The initial event is their first post with the ILI-related mention. For the control group, we picked individuals that made an ILI-related mention after the −15 ≤ *t* ≤ −2, then we picked a random post of them as an initial event in weeks −15 ≤ *t* ≤ −2, before the peak of the epidemic. Using that 30 days period, we computed different Mobility, Content, and Network traits to characterize each user.

#### Mobility traits

We worked out the mobility pattern from a user by looking at geolocations from tweets. To characterize their mobility, we used the radius of gyration^38^, which measures the size of the area covered while moving around:

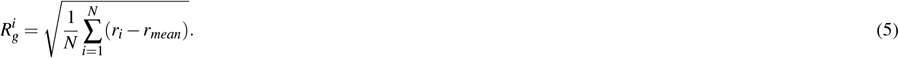

#### Content topics

We extracted topics from the texts in each user’s tweets. To this end, we use the TextRazor classifier trained against the IPTC news-codes^58^, which classify each tweet into approximately 1400 high-level categories organized into a three-level tree hierarchy. Each tweet is given a probability of containing such a topic. Thus each user is characterized by a content vector of *n* topics

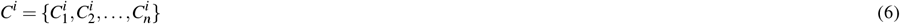

where the components 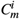 are the aggregated probability of topic *m* in all her tweets.

#### Network traits

Apart from the out-degree for each user *i* we also took into account the total user activity in the social network platform by computing the number of tweets generated during the observation period. This variable is called the number of posts.

### Linear logistic regression model

The following equation represents a linear logistic regression model for explaining the probability of an individual being a sensor by different features, where {*M*^*i*^} are the mobility features (we only consider the radius of gyration variable, *R*_*g*_), {*N*^*i*^} the group of network variables, out-degree, *d*_*out,i*_, and the number of posts, and {*C*}^*i*^ is the group of content variables for each individual *i*. Our model is

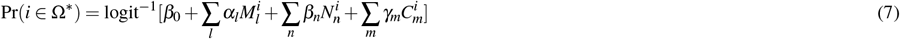

where Ω^*^ is the set of users defined as sensors, and logit^−1^(*x*) = *e*^*x*^*/*(1 + *e*^*x*^). In the model, each individual variable in the different groups is standardized to have zero mean and unit variance.

## Data Availability

All data produced are available online at

https://github.com/dmartincc/sensors

## Acknowledgements

E.M. acknowledges support by Ministerio de Ciencia e Innovación/Agencia Española de Investigación (MCIN/AEI/10.13039/501100011033) through grant PID2019-106811GB-C32. M.C. was supported by the Ministry of Universities of the Government of Spain, under the program “Convocatoria de Ayudas para la recualificación del sistema universitario español para 2021-2023, de la Universidad Carlos III de Madrid, de 1 de Julio de 2021”.

## Abbreviations

ABM=: Agent based model
EWES=: Early warning epidemiological systems
ILI=: Influenza-like illness
IPTC=: International Press Telecommunications Council
NLP=: Natural Language Processing
SIR=: Susceptible-infected-recovery

## Availability of data and materials

All data needed to evaluate the conclusions in the paper are present in the paper, the Supplementary Materials and the following repository: https://github.com/dmartincc/sensors. Additional data related to this paper may be requested from the authors.

## Competing interests

The authors have declared that no competing interests exist.

## Authors’ contributions

M.G.-H. built the code to access Twitter data from their API. D.M.-C., E.M. and M-C. designed the research. D.M.-C. performed research. D.M.-C. analysed the results. D.M.-C. and E.M. wrote the first draft of the manuscript. D.M.-C., E.M. M-C. and M.G.-H. discussed results and edited the manuscript. All authors approved the final version.

## Notes

### Competing Interest Statement

The authors have declared no competing interest.

### Funding Statement

E.M. acknowledges support by Ministerio de Ciencia e Innovacion/Agencia Espanola de Investigacion (MCIN/AEI/10.13039/501100011033) through grant PID2019-106811GB-C32. M.C. was supported by the Ministry of Universities of the Government of Spain, under the program ''Convocatoria de Ayudas para la recualificacion del sistema universitario espanol para 2021-2023, de la Universidad Carlos III de Madrid, de 1 de Julio de 2021''

### Author Declarations

The study used (or will use) ONLY openly available human data that were originally located at Twitter and Instituto Carlos III de Salud.

### Summary of Updates

Updated version of the manuscript, some changes in charts and more statistical tests.

## References

1. Parry, J. H7n9 avian flu infects humans for the first time. BMJ 346 (2013).

2. Petersen, L. R., Jamieson, D. J., Powers, A. M. & Honein, M. A. Zika virus. New Engl. J. Medicine 374, 1552–1563 (2016).

3. Stadler, K. et al. Sars—beginning to understand a new virus. Nat. Rev. Microbiol. 1, 209–218 (2003).

4. Fouchier, R. A. et al. Koch’s postulates fulfilled for sars virus. Nature 423, 240–240 (2003).

5. Feldmann, H. & Geisbert, T. W. Ebola haemorrhagic fever. The Lancet 377, 849–862 (2011).

6. Briand, S. et al. The international ebola emergency. New Engl. J. Medicine 371, 1180–1183 (2014).

7. Riou, J. & Althaus, C. L. Pattern of early human-to-human transmission of wuhan 2019 novel coronavirus (2019-ncov), december 2019 to january 2020. Eurosurveillance 25, 2000058 (2020).

8. Li, Q. et al. Early transmission dynamics in wuhan, china, of novel coronavirus–infected pneumonia. New Engl. J. Medicine (2020).

9. Pastor-Satorras, R. & Vespignani, A. Epidemic spreading in scale-free networks. Phys. review letters 86, 3200 (2001).

10. Kogan, N. E. et al. An early warning approach to monitor covid-19 activity with multiple digital traces in near real time. Sci. Adv. 7, eabd6989 (2021).

11. Ginsberg, J. et al. Detecting influenza epidemics using search engine query data. Nature 457, 1012–1014 (2009).

12. Yuan, Q. et al. Monitoring influenza epidemics in china with search query from baidu. PloS one 8, e64323 (2013).

13. McIver, D. J. & Brownstein, J. S. Wikipedia usage estimates prevalence of influenza-like illness in the united states in near real-time. PLoS Comput. Biol 10, e1003581 (2014).

14. Generous, N., Fairchild, G., Deshpande, A., Del Valle, S. Y. & Priedhorsky, R. Global disease monitoring and forecasting with wikipedia. PLoS Comput. Biol 10, e1003892 (2014).

15. Lampos, V. et al. Tracking covid-19 using online search. NPJ digital medicine 4, 1–11 (2021).

16. Soebiyanto, R. P., Adimi, F. & Kiang, R. K. Modeling and predicting seasonal influenza transmission in warm regions using climatological parameters. PloS one 5, e9450 (2010).

17. Culotta, A. Towards detecting influenza epidemics by analyzing twitter messages. In Proceedings of the first workshop on social media analytics, 115–122 (ACM, 2010).

18. Santillana, M. et al. Combining search, social media, and traditional data sources to improve influenza surveillance. PLoS Comput. Biol 11, e1004513 (2015).

19. Chen, L., Hossain, K. T., Butler, P., Ramakrishnan, N. & Prakash, B. A. Flu gone viral: Syndromic surveillance of flu on twitter using temporal topic models. In 2014 IEEE international conference on data mining, 755–760 (IEEE, 2014).

20. Ma, L.-l., Ma, C., Zhang, H.-F. & Wang, B.-H. Identifying influential spreaders in complex networks based on gravity formula. Phys. A: Stat. Mech. its Appl. 451, 205–212 (2016).

21. Christley, R. M. et al. Infection in social networks: using network analysis to identify high-risk individuals. Am. journal epidemiology 162, 1024–1031 (2005).

22. Alexander, M., Forastiere, L., Gupta, S. & Christakis, N. A. Algorithms for seeding social networks can enhance the adoption of a public health intervention in urban india. Proc. Natl. Acad. Sci. 119, e2120742119 (2022).

23. Aleta, A. et al. Quantifying the importance and location of sars-cov-2 transmission events in large metropolitan areas. Proc. Natl. Acad. Sci. 119, e2112182119 (2022).

24. Wang, Z. et al. Statistical physics of vaccination. Phys. Reports 664, 1–113 (2016).

25. Galesic, M. et al. Human social sensing is an untapped resource for computational social science. Nature 595, 214–222 (2021).

26. Ghosh, R., Mareček, J., Griggs, W. M., Souza, M. & Shorten, R. N. Predictability and fairness in social sensing. IEEE Internet Things J. (2021).

27. Rashid, M. T. & Wang, D. Covidsens: a vision on reliable social sensing for covid-19. Artif. intelligence review 54, 1–25 (2021).

28. Hodas, N. O., Kooti, F. & Lerman, K. Friendship paradox redux: Your friends are more interesting than you. ICWSM 13, 8–10 (2013).

29. Christakis, N. A. & Fowler, J. H. Social network sensors for early detection of contagious outbreaks. PloS one 5, e12948 (2010).

30. Farrahi, K., Emonet, R. & Cebrian, M. Predicting a community’s flu dynamics with mobile phone data. In Proceedings of the 18th ACM Conference on Computer Supported Cooperative Work & Social Computing, 1214–1221 (ACM, 2015).

31. Shao, H. et al. Forecasting the flu: designing social network sensors for epidemics. arXiv preprint arXiv:1602.06866 (2016).

32. Kianersi, S., Ahn, Y.-Y. & Rosenberg, M. Association between sampling method and covid-19 test positivity among undergraduate students: Testing friendship paradox in covid-19 network of transmission. medRxiv (2020).

33. Garcia-Herranz, M., Moro, E., Cebrian, M., Christakis, N. A. & Fowler, J. H. Using friends as sensors to detect global-scale contagious outbreaks. PloS one 9, e92413 (2014).

34. Dunbar, R. I., Arnaboldi, V., Conti, M. & Passarella, A. The structure of online social networks mirrors those in the offline world. Soc. networks 43, 39–47 (2015).

35. Zhang, J. & Centola, D. Social networks and health: New developments in diffusion, online and offline. Annu. Rev. Sociol. 45, 91–109 (2019).

36. Grupo de Vigilancia de Gripe del Centro Nacional de Epidemiología. Instituto de Salud Carlos III. Sistema de Vigilancia de la Gripe en España. http://vgripe.isciii.es/gripe/inicio.do. Accessed: 22-06-2019.

37. Commission, E. Commission implementing decision (eu) 2018/945 of 22 june 2018 on the communicable diseases and related special health issues to be covered by epidemiological surveillance as well as relevant case definitions. Off J Eur Union 61, 1–74 (2018).

38. González, M. C., Hidalgo, C. A. & Barabási, A.-L. Understanding individual human mobility patterns. Nature 453, 779–782 (2008).

39. Kishore, N. et al. Evaluating the reliability of mobility metrics from aggregated mobile phone data as proxies for SARS-CoV-2 transmission in the USA: a population-based study. The Lancet Digit. Heal. 4, e27–e36 (2022).

40. Preoţiuc-Pietro, D., Volkova, S., Lampos, V., Bachrach, Y. & Aletras, N. Studying User Income through Language, Behaviour and Affect in Social Media. PLoS ONE 10, e0138717 (2015).

41. Nelson, K. N. et al. Nationally representative social contact patterns among U.S. adults, August 2020-April 2021. Epidemics 40, 100605 (2022).

42. Golbeck, J., Robles, C., Edmondson, M. & Turner, K. Predicting personality from twitter. In 2011 IEEE third international conference on privacy, security, risk and trust and 2011 IEEE third international conference on social computing, 149–156 (IEEE, 2011).

43. Chamorro-Premuzic, T. & Furnham, A. Personality and music: Can traits explain how people use music in everyday life? Br. journal psychology 98, 175–185 (2007).

44. Reich, S. M., Subrahmanyam, K. & Espinoza, G. Friending, iming, and hanging out face-to-face: overlap in adolescents’ online and offline social networks. Dev. psychology 48, 356 (2012).

45. Huang, G. C. et al. Peer influences: the impact of online and offline friendship networks on adolescent smoking and alcohol use. J. Adolesc. Heal. 54, 508–514 (2014).

46. Aleta, A. et al. Modelling the impact of testing, contact tracing and household quarantine on second waves of covid-19. Nat. Hum. Behav. 4, 964–971 (2020).

47. Li, C. et al. Retrospective analysis of the possibility of predicting the covid-19 outbreak from internet searches and social media data, china, 2020. Eurosurveillance 25, 2000199 (2020).

48. Qin, L. et al. Prediction of number of cases of 2019 novel coronavirus (covid-19) using social media search index. Int. journal environmental research public health 17, 2365 (2020).

49. World Health Organization. Global outbreak alert and response network (goarn). https://extranet.who.int/goarn/.

50. World Health Organization. Integrated outbreak analytics (ioa). https://extranet.who.int/goarn/content/integrated-outbreak-analytics-delivers-holistic-understanding-outbreak-dynamics.

51. World Health Organization. Epidemic intelligence from open sources (eios). https://www.who.int/initiatives/eios.

52. World Health Organization. Epi-brain. https://www.epi-brain.com/.

53. Carter, S. E. et al. What questions we should be asking about covid-19 in humanitarian settings: perspectives from the social sciences analysis cell in the democratic republic of the congo. BMJ global health 5, e003607 (2020).

54. Twitter. Twitter developer documentation. https://dev.twitter.com/streaming/overview.

55. Pedregosa, F. et al. Scikit-learn: Machine learning in Python. J. Mach. Learn. Res. 12, 2825–2830 (2011).

56. Albert, R. & Barabási, A.-L. Statistical mechanics of complex networks. Rev. modern physics 74, 47 (2002).

57. Csardi, G. & Nepusz, T. The igraph software package for complex network research. InterJournal Complex Systems, 1695 (2006).

58. Troncy, R. Bringing the iptc news architecture into the semantic web. In International Semantic Web Conference, 483–498 (Springer, 2008).

