## Supplementary Materials for "Social Media Sensors to Detect Early Warnings of Influenza at Scale"

#### Supplementary Sections

**1 Data** 2

**2 Centrality sensitivity analysis** 2

**3 Sensors selection analysis** 3

**4 Agent-based model of ILI disease and information diffusion** 3

**5 Sensors logistic regression model** 4

### 1 Data

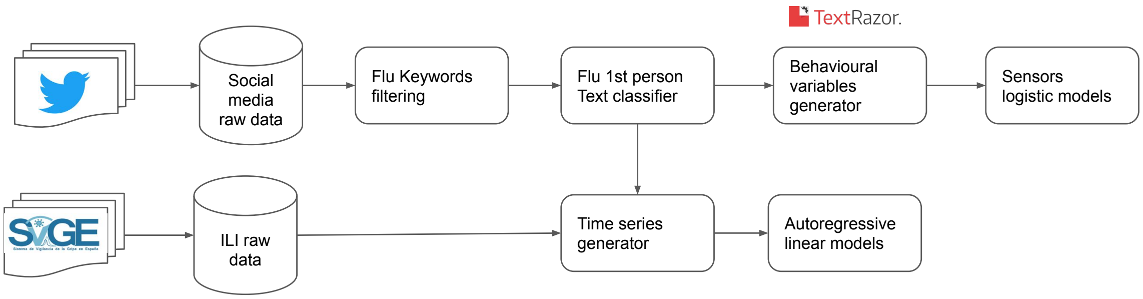


**Figure S1: Data processing pipeline schematic view.** From left to right, raw data collectors, data filtering, data enrichment, time series generator, and models.

In Figure S1 we can see our data processing pipeline is composed of three different stages. First, data was accessed and analyzed from the Twitter API and the surveillance system for influenza in Spain (ScVGE) [1]. Second, data cleaning. This stage has an ILI keyword-based filtering step, where we look for ILI-related Spanish words, such as ”gripe”, ”gripazo”, ”trancazo”, ”catarro” and ”constipado”, and a second step with a first-person ILI-related posts text classifier. Then our pipeline is divided into two branches. One for building autoregressive linear models for explaining and predicting official weekly ILI cases, where is required a previous step for grouping by weeks the transactional data before feeding our linear models. The second branch builds several logistic linear models at the individual level for explaining and predicting potential sensors from the network. Before feeding the models, we create some behavioral variables, based on social activity, mobility, and content posting.

### 2 Centrality sensitivity analysis

Figure S2 shows the time series of different centrality metrics of the users making ILI-related posts compared with the total number of them. We show the average during the 2013, 2014, and 2015 seasons of ILI in Spain. Time series are centered around their maximum peak within the season. For the centrality metrics, we show the weekly total out-degree ($D_{T,t}$ in the main text) compared with the weekly average outdegree ($D_{T,t}$ divided over the number of users making ILI-related posts) and the weekly median of the out-degree of users making ILI-related posts. As we can see, the total out-degree clearly follows the number of ILI-related mentions and shows a large spike weeks before the peak. That peak is also observed for the average degree. However, it is not present in the median. These results show that high-connected people have ILI-related posts on the social network at different times than others. And it only appears in the total or average degree since those estimators are more susceptible to large out-degree users than the median out-degree.

To make that difference more quantitative in Figure S3 we show the difference on the average of those centrality metrics 18 weeks before and after the peak. We applied the statistical test t-test to see if there were differences between the groups. Figure S3. As we can see, the total out-degree is the one that shows clearly more difference before and after the peak. For that reason, we choose it in the main paper.

#### 3 Sensors selection analysis

To define sensors, Figure S4 shows the out-degree distribution of all the users having ILI mentions. Again, it follows a power-law distribution with an exponent of 2.56 (CI $[2.51, 2.62]$). Based on this distribution, we defined four out-degree thresholds to test different groups of sensors. Out of the users making ILI-related mentions, we defined a sensor as a user with an out-degree greater than 100, 500, 1000, or 2000 (vertical dashed lines in Figure S4), and as control otherwise. Figures S4B shows the results for the cumulative incidence of the ILI-related mentions for each of the out-degree sensor thresholds. As we can see, for an out-degree threshold greater than 1000, the cumulative ILI-related mentions incidence for the sensor group is ahead and starts to grow one or two weeks before the control group. Therefore, we selected the out-degree threshold to be 1000 from now onward in our study.

#### 4 Agent-based model of ILI disease and information diffusion

The values of all the parameters used in our Agent-Based Model (ABM) simulating the Susceptible-Infected-Recovery epidemic spreading on a complex network are given in Table S1. The synthetic network is generated by the Barabasi-Albert model using the `igraph` R package [2]. We simulated different realizations of the epidemic model, see Figure S5, emulating different ILI seasons. To compare the temporal dynamics of our ABM with the real ILI epidemics, we rescale the time in our model to mimic the epidemic dynamics in the empirical data. As we can see in Figure S5 equating four-time units in our simulations to one week, the epidemic curves have the same shape as the real ILI-related cases.

We also assume that each agent posts on a social media platform and that those tweets are ILI-related when he gets infected. To incorporate our hypothesis that offline and online networks degrees are correlated, we assume that

$$d_i^{\text{Twitter}} = d_i^{\text{Offline}}(1 + \nu_i)$$

where $\nu_i$ is a random number uniformly distributed between 0 and 1. This way, we account for potential variability between offline and online degrees while still getting a moderate correlation between them.

In this ABM model, we assume that sensors in the social media platform are those with $d_i^{Twitter} \geq \chi$, see Table S1. Figure S5C shows the average total out-degree for all users and those in the sensor group compared with the number of infected agents. As expected, users with a larger degree get infected earlier and those in the sensor group even a little bit earlier, as we saw in the real data.

| Parameters | Description | Value |
|---|---|---|
| $N$ | Number of nodes in graph | 150k |
| $n$ | Initial seeds | 2 |
| $\beta$ | Infection probability ($S \to I$) | 10% [3] |
| $\epsilon$ | Latent period | 3 days [3] |
| $\alpha$ | Recovery probability ($I \to R$) | 1 / $\epsilon$ |
| $\chi$ | Sensors degree threshold | 12 |

**Table S1: SIR Agent-based model parameters.** Set of parameters used in our agent-based model of ILI disease and information propagation.

### 5 Sensors logistic regression model

In table S2, we can see the coefficients of the logistic regression models to explain and identify a single node as a sensor. We used three groups of variables, a categorization of the content published, the user's mobility, and user's network features, including their out-degree and number of posts.

| | *Dependent variable:* | | | |
|---|---|---|---|---|
| | Sensor | | | |
| | Content | Network | Mobility | All |
| | (1) | (2) | (3) | (4) |
| Association | $-0.263^{***}$ (0.099) | | | |
| Basketball | | | | $-0.237^{**}$ (0.098) |
| Christianity | $0.116^{**}$ (0.053) | | | 0.082 (0.054) |
| Christmas | $0.134^{**}$ (0.060) | | | 0.097 (0.061) |
| Easter | $-0.144^{**}$ (0.066) | | | $-0.133^{**}$ (0.067) |
| Entertainment | $-0.154^{*}$ (0.093) | | | $-0.164^{*}$ (0.094) |
| Folk | $-0.237^{***}$ (0.080) | | | $-0.225^{***}$ (0.081) |
| Government | 0.125 (0.081) | | | $0.189^{***}$ (0.058) |
| Human | $0.118^{**}$ (0.058) | | | $0.103^{*}$ (0.058) |
| Language | $0.218^{***}$ (0.057) | | | $0.156^{***}$ (0.058) |
| Music | $0.499^{***}$ (0.127) | | | $0.461^{***}$ (0.129) |
| National | $0.292^{***}$ (0.113) | | | $0.168^{*}$ (0.093) |
| Organisations | $0.156^{**}$ (0.074) | | | 0.109 (0.072) |
| Philosophical | 0.090 (0.057) | | | |
| Politics | $0.133^{*}$ (0.081) | | | |
| Popular | $-0.160^{**}$ (0.076) | | | $-0.154^{**}$ (0.077) |
| Soccer | $-0.138$ (0.087) | | | |
| Out-degree | | $0.549^{**}$ (0.233) | | $0.511^{**}$ (0.228) |
| Number of posts | | $0.461^{***}$ (0.073) | | $0.315^{***}$ (0.074) |
| Radius of gyration | | | $0.113^{**}$ (0.053) | $0.104^{*}$ (0.056) |
| Constant | $-0.492^{***}$ (0.056) | $-0.444^{***}$ (0.055) | $-0.463^{***}$ (0.054) | $-0.470^{***}$ (0.057) |
| Observations | 1,460 | 1,460 | 1,460 | 1,460 |
| Accuracy | 0.605 | 0.607 | 0.615 | 0.636 |
| Accuracy CI | (0.568, 0.632) | (0.567, 0.645) | (0.575, 0.653) | (0.596, 0.673) |
| Log Likelihood | −926.504 | −941.322 | −971.694 | −913.783 |
| Akaike Inf. Crit. | 1,887.008 | 1,888.645 | 1,947.388 | 1,861.566 |
| *Note:* | | | | $^{*}$p<0.1; $^{**}$p<0.05; $^{***}$p<0.01 |

**Table S2: Sensor models.** Logistic regression models for sensors characterization based on content, network, and mobility features.

### References


[1] Grupo de Vigilancia de Gripe del Centro Nacional de Epidemiologa. Instituto de Salud Carlos III: Sistema de Vigilancia de la Gripe en Espaa. http://vgripe.isciii.es/gripe/inicio.do. Accessed: 22-06-2019

[2] Csardi, G., Nepusz, T.: The igraph software package for complex network research. InterJournal **Complex Systems**, 1695 (2006)

[3] Tokars, J.I., Olsen, S.J., Reed, C.: Seasonal incidence of symptomatic influenza in the united states. Clinical Infectious Diseases **66**(10), 1511–1518 (2018)

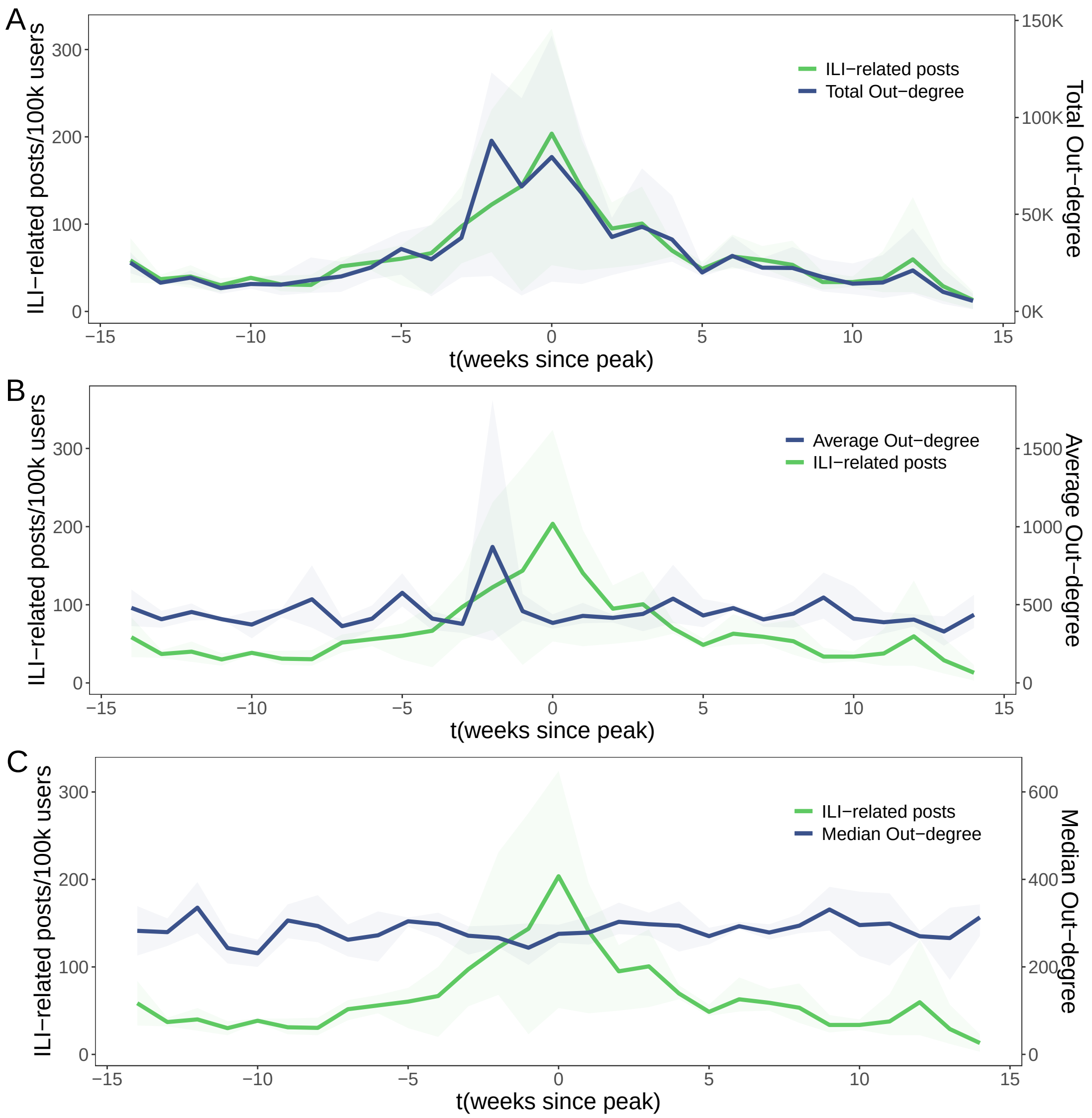


**Figure S2: Generalized ILI-related posts compared with weekly centrality statistics in Twitter.** Horizontal axis measures weeks from the peak. Green solid lines show the average incidence across seasons of weekly ILI-related posts (left Y-axis). Blue lines represent the average different weekly centrality metrics from individuals posting a first-person ILI-related post (right Y-axis). (A) is the weekly total out-degree, (B) is their average out-degree, and (C) is the median out-degree of those individuals. Shaded areas are the confidence intervals over the different seasons.

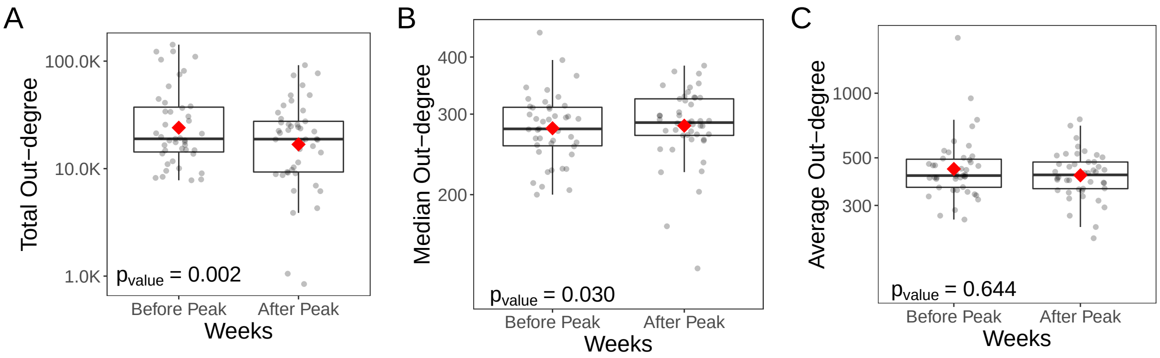


**Figure S3: Comparison of degree metrics before and after peak groups.** Points correspond to different weeks grouped by before and after the peak. We also show the box-plot for each group, including their median (horizontal thick line) and mean (red diamonds). Vertical axis are centrality metrics: (A) Total out-degree, (B) Median out-degree and (C) Average out-degree. The p-value shows the t-test statistic comparing the means for the groups before and after the peak.

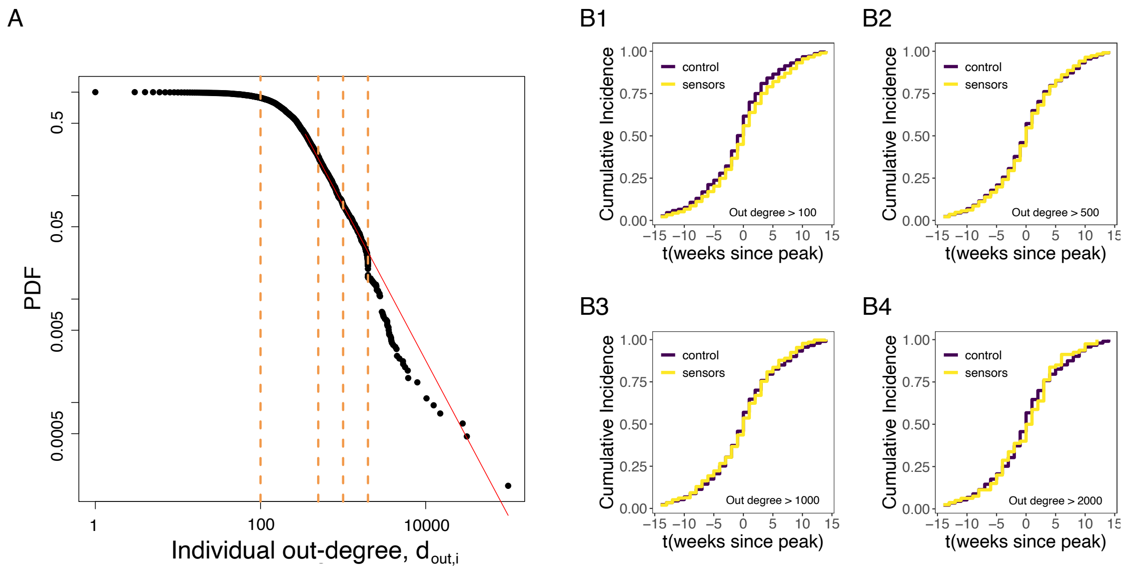


**Figure S4: Out-degree sensitivity analysis.**(A) Users out-degree $d_{out,i}$ frequency distribution. Vertical orange dashed lines define the thresholds selected with out-degrees 100, 500, 1000, and 2000. (B) Empirical cumulative distribution differences in ILI-related mentions on Twitter between the sensor and randomly chosen individuals for each of the degree thresholds selected. The purple line correspond to the group of randomly chosen individuals and the yellow line is sensor group selected with an out-degree bigger than 100 (B1), 500 (B2), 1000 (B3) and 2000 (B4).

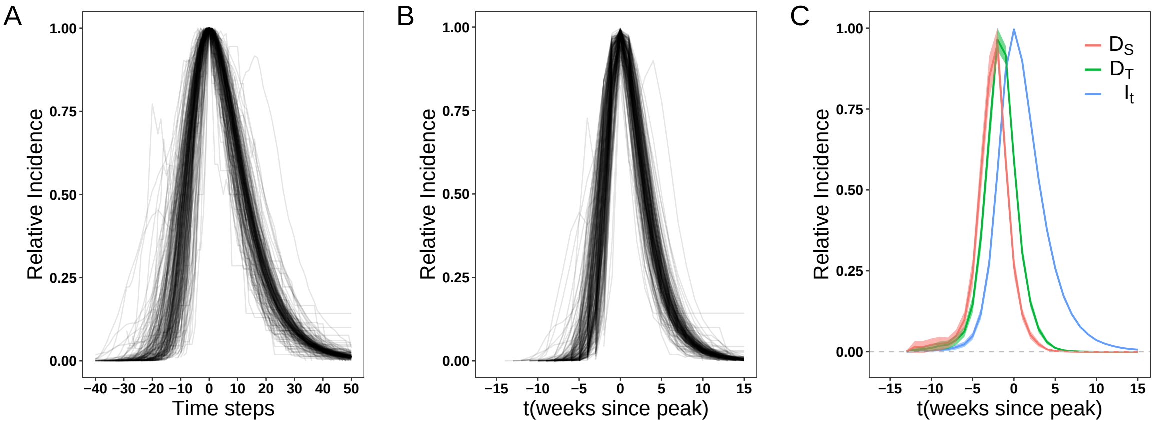


**Figure S5: Agent-based simulations of ILI disease and information diffusion.** (A) Shows the incidence curves for the spreading of the diseases for each simulation in our original time scale. Time is centered around the peak for each simulation, and we show the relative incidence to its maximum (peak). (B) Same as in A), but with time steps converted into weeks to compare with real ILI cases. (C) Average total out-degree in the social network for all agents ($D_T$) and those in the sensor group ($D_S$) compared with the incidence of the disease. For comparison, out-degree is normalized to the maximum in each curve.
